# Virtual Delivery of Simulation Education to Undergraduate Medical Students During the COVID-19 Pandemic

**DOI:** 10.1101/2021.08.20.21262347

**Authors:** Kay Wu, Alex Chan, Avinash Pandey, Puru Panchal, Maroof Khalid, Sudarshan Bala, Samveg Shah, Matthew C. Miller

## Abstract

**Background:** The COVID-19 pandemic has restricted in-person clinical training for medical students. Simulation-based teaching is a promising tool to introduce learners to the clinical environment. MacSim is a student-led simulation workshop for learners to develop clinical competencies. The objective of this study was to assess the impacts of MacSim and participants’ perspectives regarding simulation-based teaching.

**Methods:** A comprehensive simulation, representative of a virtual care scenario, was delivered to 42 pre-clerkship medical students via video conferencing. In pairs, participants obtained histories and carried out management plans for simulated patients. Participants were surveyed and interviewed. Survey data were analyzed using the Wilcoxon signed-ranks test. Interview transcript data were thematically analyzed.

**Results:** Post-simulation, participants (n=24) felt more prepared to make clinical decisions, collaborate, and communicate in a virtual setting. 92% of respondents agreed MacSim was a valuable learning experience and 96% agreed more simulation-based learning should be integrated into curricula. Emergent themes from interviews (n=12) included: 1) value of simulation fidelity, 2) value of physician feedback, and 3) effectiveness of MacSim in improving virtual clinical skills.

**Conclusion:** Simulation-based teaching is of importance and educational value to medical students. It may play an increasingly prevalent role in education as virtual care is likely to become more prevalent.

## Introduction

To adhere to social distancing guidelines introduced by the COVID-19 pandemic, a large proportion of educational activities have migrated to e-learning programs, and clinical encounters to telehealth provision. ^1–3^ Though these changes have provided benefits in allowing healthcare providers to continue providing care, its introduction brings new challenges to practitioners and learners as new dimensions and barriers are added to patient interaction.^4,5^ Though novel strategies have been adopted to adapt to these changes, the introduction of novel clinical practices nonetheless bring on new challenges to developing pre-clerks. ^4-7^ A promising tool being used to introduce pre-clerk students into the virtual clinic environment is simulation-based teaching. Traditionally, simulation-based teaching has been used at all levels of medical education, providing students a risk-free environment to gain necessary competencies to be proficient in the clinical environment.^8–12^

Though there have been extensive studies examining the efficacy of simulation-based teaching in different settings, few studies have characterized the efficacy of simulation teaching as a tool for integrating medical students into the virtual clinic environment, and the inclusion of simulation-based teaching can often be sparse for junior learners. ^13^ The present study sought to address this knowledge gap by investigating perspectives and potential changes in self-perceived competencies of medical students after a virtual simulation.

## Methods

### Study Design

This cross-sectional study consisted of two components centred around a simulation event. The simulation event consisting of a 30-minute session in which participants had 20 minutes with a simulated patient in a virtual clinic environment, followed by 10 minutes of feedback from a physician assessor. The study design included comparison of pre- and post-event survey data, as well as post-event interview to assess perspectives regarding simulation teaching and COVID-19.^14^ The study was PIPEDA-compliant and exempted from ethics approval by the Hamilton Integrated Research Ethics Board due to it being a quality improvement project.

### Recruitment

Advertising of the event was done internally via social media and e-mail at the host school to second year students two weeks prior to the event. No financial incentives were included, and forty-two attendees participated in the event.

### Survey and Interview Design

An online survey was sent to the attendees prior to and directly after the event. Non-identifying demographic data were obtained. Four Likert scale items were used to gauge perception of preparedness for clinical duties in the virtual environment, including history taking, clinical decision making, working in a team, and communicating with patients, measured on a scale of 1 (not at all prepared) to 5 (very well prepared). Data on participant perceptions on simulation learning were additionally gathered via Likert scales ranging from 1 (strongly disagree) to 5 (strongly agree). Statements used in the study to gauge student perception on the value of simulation-based learning included items such as “This event was a valuable learning experience to further familiarize myself with virtual clinics”, “More simulation-based learning should be integrated into the pre-clerkship curriculum”, and “COVID-19 has significantly reduced my opportunities for learning in a clinical setting”.

After the event, one-on-one interviews were conducted and transcribed verbatim with 12 randomly selected participants by one of the authors. Interviews lasted 20-30 minutes, utilizing the Modified Simulation Effectiveness Tool (SET-M), a validated tool for evaluation of simulation exercises in both nursing and medical students.^14^

### Statistical Analysis

Analysis was done using Statistical Package for the Social Sciences (SPSS) 26. The Wilcoxon Signed-Rank Test was used to compare survey responses obtained before and immediately after the event following testing for parametric assumptions.^15-16^ Likert scale data were compared between pre- and post-event surveys to determine if there was an immediate impact in self-perceived preparedness for integration into the virtual clinic environment.

### Qualitative Analysis

Analysis of the interviews was performed according to the six-step process described by Braun and Clarke.^17^ Interview transcripts were independently coded by two authors. Transcript data and codes were then qualitatively analyzed by the team to reach a consensus on relevant themes and subthemes.

## Results

### Comparison of Survey Data

There were 28 (70% response rate) and 24 responses (60% response rate) for the surveys sent before and immediately after the survey, respectively. The average age of attendees was 23.3 years (SD = 2.2). The post-event survey showed significantly higher scores of perception of preparedness for history taking in the virtual clinic environment [pre-event Mdn = 4 (SD = 0.63), post-event Mdn = 4 (SD = 0.48), Z = 2.13, *p* < 0.05], preparedness in clinical decision making [pre-event Mdn = 3 (SD = 0.57), post-event Mdn = 3 (SD = 0.79), Z = 3.35, *p* = 0.001], working in a team setting [pre-event Mdn = 3 (SD = 0.79), post-event Mdn = 4 (SD = 0.77), Z = 2.51, *p* <.05], and communicating to patients in a virtual clinic environment [pre-event Mdn = 3 (SD = 0.80), post-event Mdn = 4 (SD = 0.69), Z = 2.17, *p* <.05]. Furthermore, participants agreed that the event was a valuable experience for their learning (Mean = 4.29, SD = 0.62), strongly agreed that more simulation-based learning should be integrated into the pre-clerkship curriculum (Mean = 4.58, SD = 0.58), and strongly agreed that COVID-19 has significantly reduced their opportunities for learning in a clinical environment (Mean = 4.83, SD = 0.48). Results are summarized in Figure 1.

**Figure 1:**
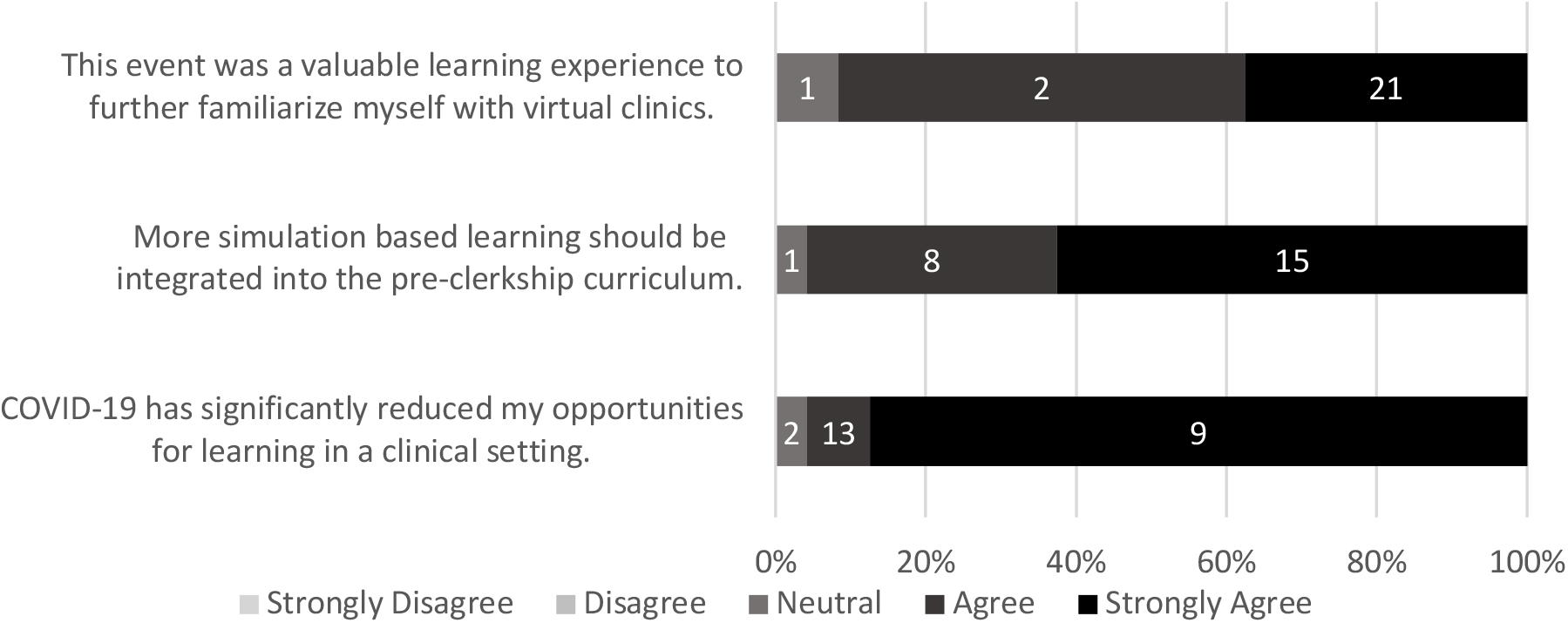
Likert Scale Data Obtained from the Post-Event Survey

### Analysis of Interview Data

Five major thematic categories emerged from the qualitative analysis (Table 1).

**Table 1:** Themes Identified upon Qualitative Analysis and Sample Quotes from Participant Interviews

| **Theme** | **Percentage of Respondents Who Identified Theme** | **Example Quotes** |
| --- | --- | --- |
| **Collaboration and Communication** | 9/12 | My partner and I actually live together, so we did it side by side, so we could talk and make decisions together.<br>I thought that [working in pairs] was really helpful because we were able to bounce ideas off of each other in person.<br>Our resident asked us [each] to do a brief one-liner/HPI of the patient.<br>Liked working with one partner. |
| **Simulation Fidelity** | 9/12 | The consult note was very extensive and good practice for clerkship for combing through a dense file for what you really need.<br>The way SP presented and weren't very sure about their history and didn't give precise answers made you better at probing and finding little tricks like different |
|  |  | ways to ask questions, because it's not like you'll always get the perfect answer.<br>The fact that the case wasn't too complicated where I felt overwhelmed about what to do: it was simple enough that at our level, we should realistically know what the next steps would be with relative confidence. This was the best thing for the learning. |
| **Virtual Simulation Advantages and Disadvantages** | 8/12 | We got some helpful tips from our resident just about maneuvers, like getting a family member to do a physical exam—that sort of stuff that I didn't think of before.<br>Given the sim was online, I thought it was pretty realistic. I thought it was good that it included lots of different aspects, like history, physical exam, and investigations, which I think is pretty difficult to do so I thought that was pretty good.<br>I think the method of asking for an investigation and you guys posting the results was actually really smart and went really well.<br>I enjoyed the "fast-forwarded" nature of the patient care experience. I liked having tests available quickly.<br>I can see how if it was over Zoom and you weren't next to each other it could be more awkward, but I liked that partner aspect. |
| **Knowledge and Skill Development** | 10/12 | Will remember diagnostic pathways more having thought through them compared to reading in textbook.<br>Awesome that you guys didn't just give us labs, but gave more prompting so that we had to decide which labs are relevant.<br>We had to decide whether or not the case was urgent and we also had to decide what to do for the patient as they present so that was a good learning opportunity.<br>[Our evaluator] was very good about explaining the details and directing us on how to effectively and efficiently gain information to present to our attendings. It was useful to recap why we made certain decisions and understand there's a disconnect between textbook presentation of a case and how someone might present.<br>The sim did stimulate me to look up the pathophysiology after, so I do have a better understanding now. |
| **Suggestions for Use in Undergraduate Medical Education** | 11/12 | I think if MacSim could be integrated into the program somehow, it would be helpful. I think this is more valuable more near the end of pre-clerkship, or even having one sim at the end of each foundation, because it gets you more in the clinical mindset instead of just the pathophysiology and history taking. |
|  |  | It would be nice to have MacSim, especially approaching clerkship. With [Transition to Clerkship] electives it was variable how much autonomy I had. I would say a few sessions, maybe two or three, would be nice to help us practice in some basic cases. I think a number of sim cases that are at our level and allow us to make decisions would allow us to get familiar with communicating those decisions to patients outside of a formal OSCE and clinical skills setting.<br>I wish we did more of this at the end of each foundation as review. The trouble with tutorials is that they give you the case and they're very classical presentations or a very stereotypical picture, so it's easy to know what the diagnosis is because they spell it out for you. That's also the situation with our OSCE-type scenarios. As we're starting clinical rotations or pre-clerkship electives we're starting to notice gathering information is very difficult—it's not always spelled out for you, so I think this type of real life situation is very useful. |

The “Collaboration and Communication” theme reflected the value of the exercise for learning how to work with a partner to make clinical decisions, for discussing and evaluating ideas, and for practicing case presentations.

The “Simulation Fidelity” theme revealed the value of simulation that was reflective of reality. One participant commented on the relatively less stressful nature of the simulation compared to a real case.

According to most participants, the simulation was effective for gaining practice for conducting histories and physical examinations virtually. Some participants commented on the challenges of conversing about decision-making through an online format in the presence of the standardized patient.

Multiple participants identified that the simulation exercise helped with knowledge and skill development and identified feedback sessions with adjudicators to be particularly useful.

Most participants stated that they would be in favor of integration of such simulation exercises into the pre-clerkship curriculum. Participants identified that it would be useful at multiple timepoints for knowledge integration and confidence-building prior to the start of clinical rotations.

## Discussion

There is a paucity of information that examines the effectiveness of simulation-based teaching in the virtual clinical setting, and how attitudes toward simulation-based teaching may have changed during the COVID-19 pandemic.

Comparison of Likert Scale data from pre- and post-event surveys indicated statistically significant differences suggesting that participants felt significantly more prepared in all areas of history taking, working in a team, communicating with patients, and making clinical decisions. Though few studies have directly compared pre- and post-intervention data, this is in opposition to a prior study that found no differences in observable clinical outcomes when comparing the pre- and post-intervention groups after one extended simulation session, possibly demonstrating a discrepancy for when subjective and objective differences occur post-intervention.^18^

In our study, thematic analysis of interview questions showed similar themes to published qualitative analyses of medical simulation. A 2006 study of clerkship students interacting with robot simulators noted that high-fidelity simulation made students “feel [they] are interacting with a live patient”^18(p217)^. This study also noted a theme, “suggestions for use and place in undergrad medical education”, where as much as 22% of participants suggested more frequent and mandatory simulations in clerkship.^18^ During the COVID-19 pandemic, where virtual encounters are becoming the norm for outpatient visits, participants in our study were also noted to frequently compliment the fidelity of our simulation and request more frequent and mandatory virtual simulations in pre-clerkship. Several other studies have also noted themes of knowledge and skill development with simulation learning.^19^ A 2020 mixed-methods study of simulated scenario teaching run by peers also commented extensively on the value of peer feedback and observing peers participating in simulated encounters.^19^ It was noted that in review of the literature, previous studies with either quantitative or qualitative analysis of simulation of virtual interviews was not found.

Several limitations are recognized for the present study. Sampling bias and nonresponse bias may have factored into the survey data of the present study as participants involved in the study were solely composed of participants in the simulation event. A lack of a validated survey for data collection, and a small sample size may additionally impact the validity of findings.

## Conclusion

To our knowledge, this study is the first mixed-methods evaluation of a student-run virtual care simulation for undergraduate medical students in the COVID-19 era. Data from our study provides new evidence to suggest that simulation-based learning may be a promising tool for improving student competencies in the virtual clinical setting. With further validation, these findings may demonstrate a need for undergraduate medical curricula to place a higher emphasis on simulation-based teaching as virtual clinics will continue to occupy a prominent role in medical practice.

## Data Availability

There is no additional data made available.

